# Brain-Derived pTau217 and Plasma Proteomic Profiles Across Cognitive and Dementia Groups in Congolese Older Adults

**DOI:** 10.64898/2026.10.26.26366533

**Authors:** Jean Ikanga, Saranya Sundaram Patel, Theresa Boots, Xuemei Zeng, Caterina Obenauf, Guy Gikelekele, Emmanuel Epenge, Japhet Magolu Potshi, Topina Tomadia, Francine Manyonga Sabowa, Joseph Phuati Tsangu, Samuel Mampunza, Lelo Mananga, Alvaro Alonso, Alden L. Gross, Thomas Karikari

**Affiliations:** Emory University School of Medicine, Department of Rehabilitation Medicine, Atlanta, GA 30322, USA; OneRehab, Dallas, Texas, USA; University of Pittsburgh, Department of Psychiatry, School of Medicine, PA, USA; University of Tennessee, Department of Psychology and Neuroscience, Knoxville, TN, USA; University of Kinshasa and Catholic University of Congo, School of Medicine, Kinshasa, Department of Psychiatry, B.P. 7463 Kinshasa I, Democratic Republic of Congo; University of Kinshasa, Department of Neurology, Kinshasa, B.P. 7463 Kinshasa I, Democratic Republic of Congo; Memory Clinic of Kinshasa, Kinshasa, B.P. 7463 Kinshasa I, Democratic Republic of Congo; University of Kinshasa, Department of Anesthesiology and Reanimation, B.P. 7463 Kinshasa, Democratic Republic of Congo; Department of Psychiatry, B.P. 7463 Kinshasa I, Democratic Republic of Congo; Department of Preventive Medicine and Public Health, School of Medicine, University of Navarra, Pamplona, Spain; Department of Epidemiology, Johns Hopkins Bloomberg School of Public Health, Baltimore, Maryland, USA

## Abstract

Blood-based biomarkers for Alzheimer disease and related dementias have been validated largely in populations of European ancestry. We characterized plasma proteins in 786 adults aged 65 years or older in Kinshasa, Democratic Republic of the Congo, classified as cognitively unimpaired, mild cognitive impairment, suspected Alzheimer disease, mixed dementia, or vascular dementia. Proteins were quantified with the NULISAseq CNS Disease Panel 120. Sex showed the strongest associations with protein expression, followed by age, whereas APOE ε4 carrier status was associated with four measures. MME was downregulated across all dementia groups and showed the largest effect in each dementia-versus-CU comparison. A random forest model combining 10 biomarkers with age, sex, and APOE ε4 carrier status achieved a mean cross-validated area under the curve of 0.768, compared with 0.612 for brain-derived pTau217 alone. Because biomarker selection and ranking were performed using the full dataset before cross-validation, model performance may be optimistic. These findings support demographic adjustment and population-specific validation of plasma biomarkers and identify MME as a hypothesis-generating signal for further study.

## INTRODUCTION

Blood-based biomarkers offer a scalable and minimally invasive approach to evaluating neurodegenerative disease compared with cerebrospinal fluid sampling or amyloid-β and tau positron emission tomography (PET), an advantage that may be particularly important in low- and middle-income countries.^1,2^ High-throughput proteomic technologies can quantify many plasma proteins simultaneously and characterize biological pathways beyond amyloid and tau pathology. These technologies include bead-based multiplex immunoassays, mass spectrometry, Olink proximity extension assays, SomaScan, and the nucleic acid-linked immuno-sandwich assay (NULISA). NULISA provides highly sensitive multiplex measurement of low-abundance proteins, including central nervous system (CNS)-enriched forms of phosphorylated tau (p-tau).^3^

The NULISAseq CNS Disease Panel 120 measures more than 120 proteins spanning amyloid and tau pathology, synaptic function, neurodegeneration, inflammation, and vascular and metabolic processes. Representative analytes include APOE, p-tau isoforms, α-synuclein (SNCA), VGF, membrane metalloendopeptidase (MME; neprilysin), vascular endothelial growth factor A (VEGFA), neurofilament light chain (NEFL), synaptosomal-associated protein 25 (SNAP25), and multiple cytokines. Studies using NULISA have identified plasma protein profiles associated with mild cognitive impairment (MCI), Alzheimer disease (AD) and related dementias, amyloid burden, and other neurodegenerative phenotypes.^4–10^ Rea Reyes and colleagues identified 19 proteins associated with clinically adjudicated MCI and reported that NULISA-measured BD-pTau217 performed comparably to Simoa-based assays for discriminating amyloid-PET status.^10^

Of particular interest is brain-derived pTau217 (BD-pTau217), a measure designed to enrich for CNS-derived pTau217 and reduce the contribution of peripheral tau. Previous studies have reported strong discrimination of AD-related amyloid pathology and larger fold differences than those observed with conventional plasma pTau217.^10,11^ BD-pTau217 may also be less susceptible to variation associated with chronic kidney disease, body mass index, hypertension, and vascular comorbidity.^11^ Combining BD-pTau217 with other proteins may provide additional information about AD-related biological changes and disease heterogeneity.^12^ These findings support BD-pTau217 as a promising component of blood-based biomarker strategies, while also highlighting the need to interpret it against the biological or clinical outcome used for validation.

Most plasma proteomic studies of dementia have been conducted in North American and European cohorts, with limited representation from African populations.^13^ Biomarker concentrations and their associations with disease may vary with genetic background, environmental exposures, vascular and metabolic comorbidities, and preanalytical factors.^14–17^ Recent work from Nigeria has begun to address this evidence gap, but data from continental Africa remain limited.^18^ Inflammatory pathways and APOE ε4 may also show different associations with cognitive decline and AD risk across African and African-ancestry populations.^13,19–21^ It therefore remains unclear whether biomarker profiles and classification approaches developed largely in high-income settings generalize to older adults in the Democratic Republic of the Congo (DRC).

We characterized plasma proteomic profiles among older adults in the DRC who were clinically classified as cognitively unimpaired (CU), MCI, suspected AD, mixed dementia, or vascular dementia. We examined associations of protein concentrations with age, sex, and APOE ε4 carrier status; identified proteins that differed across diagnostic groups; and developed random forest models to distinguish CU from suspected AD. Because amyloid PET and cerebrospinal fluid biomarkers were unavailable, the performance of BD-pTau217 was evaluated against clinical diagnosis and was not interpreted as evidence of biomarker-confirmed amyloid status. We hypothesized that demographic and genetic factors would be associated with plasma protein profiles and that multianalyte models would discriminate CU from suspected AD more accurately than BD-pTau217 alone.

## RESULTS

The analytic sample included 786 participants: 149 CU, 226 with MCI, 205 with suspected AD, 88 with mixed dementia (RD-M), and 118 with vascular dementia (RD-V). The sample included 501 women (64%) and 285 men (36%). Age and MoCA scores differed across diagnostic groups (both P < .001), as did the distribution of sex (P = .019). APOE ε4 carrier status did not differ by diagnostic group (P > .9; Table 1).

**Table 1.** Participant Characteristics by Diagnostic Group.

|  | **Total**<br>**(n = 786)** | **CU (n =**<br>**149)** | **MCI**<br>**(n = 226)** | **AD**<br>**(n = 205)** | **RD-M**<br>**(n = 88)** | **RD-V**<br>**(n = 118)** | **p-value** |
| --- | --- | --- | --- | --- | --- | --- | --- |
| Sex |  |  |  |  |  |  | 0.019 |
| *Male* | 285 (36%) | 65<br>(44%) | 84<br>(37%) | 74<br>(36%) | 34<br>(39%) | 28<br>(24%) |  |
| *Female* | 501 (64%) | 84 (56%) | 142 (63%) | 131 (64%) | 54 (61%) | 90 (76%) |  |
| Age (years) | 72 (9) | 70 (7) | 71 (8) | 74 (10) | 73 (8) | 74 (7) | < 0.001 |
| MoCA<br>Score (sum) | 20 (7.0) | 24 (4.0) | 21 (3.8) | 16 (7.0) | 17 (6.0) | 19 (6.0) | < 0.001 |
| APOE ε4<br>carrier status |  |  |  |  |  |  | > 0.9 |
| *Carrier* | 335 (43%) | 62<br>(42%) | 100<br>(44%) | 87<br>(42%) | 34<br>(39%) | 52<br>(44%) |  |
| *Non-Carrier* | 451 (57%) | 87 (58%) | 126 (56%) | 118 (58%) | 54 (61%) | 66 (56%) |  |

Note. APOE ε4 carrier status was inferred from APOE4 NPQ values using a cutoff of 5.78; values below 5.78 were classified as noncarriers, and values of 5.78 or higher were classified as carriers. Heterozygous and homozygous carriers were not distinguished.

### Associations of Demographic Variables With Biomarker Expression

Sex showed the broadest and largest associations with biomarker expression, followed by age, whereas APOE ε4 carrier status showed the fewest associations. The three largest sex-associated increases were observed for MME, HBA1, and IL13 (Figure 1). Relative to women, men had higher levels of MME (fold change [FC] = 1.49), HBA1 (FC = 1.50), and IL13 (FC = 1.27). The three largest decreases in men were observed for SAA1 (FC = 0.56), CRH (FC = 0.74), and BDNF (FC = 0.78).

**Figure 1.**
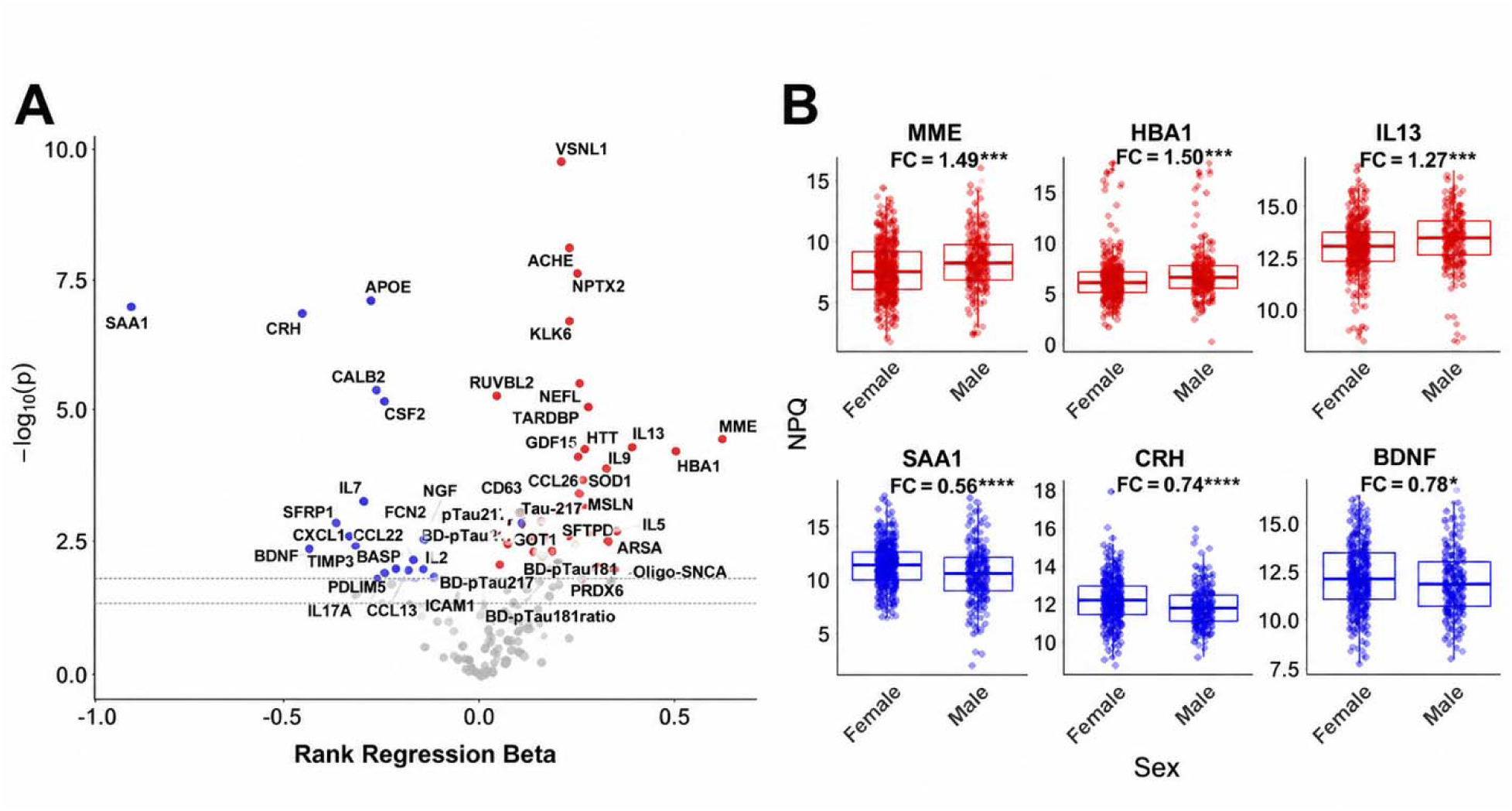
Association of sex with biomarker expression. a, Volcano plot for the full NULISA panel. P values and regression coefficients are from the sex covariate in the rank-based regression model for each biomarker. The dashed line marks the false discovery rate-adjusted significance threshold; labeled points met the adjusted P < .05 criterion. b, Distributions of the three biomarkers with the largest increases and the three biomarkers with the largest decreases in men relative to women.

Age was associated with several established neurodegeneration- and AD-related markers, including GFAP, NEFH, NEFL, BD-pTau231, and MME, although the age coefficients were generally smaller than the sex coefficients (Supplementary Figure 1a,b). Only four measures differed significantly by APOE ε4 carrier status after false discovery rate correction: pTau217 ratio, pTau231 ratio, BD-pTau217 ratio, and CRP (Supplementary Figure 1c,d).

Figure 2 compares effect estimates for sex, age, and APOE ε4 carrier status across selected AD-related biomarkers in the combined cohort and within the CU, MCI, AD, and other dementia groups. In the combined cohort, sex generally showed larger effect estimates than age, although the magnitude and direction of these associations varied by biomarker and diagnostic group.

**Figure 2.**
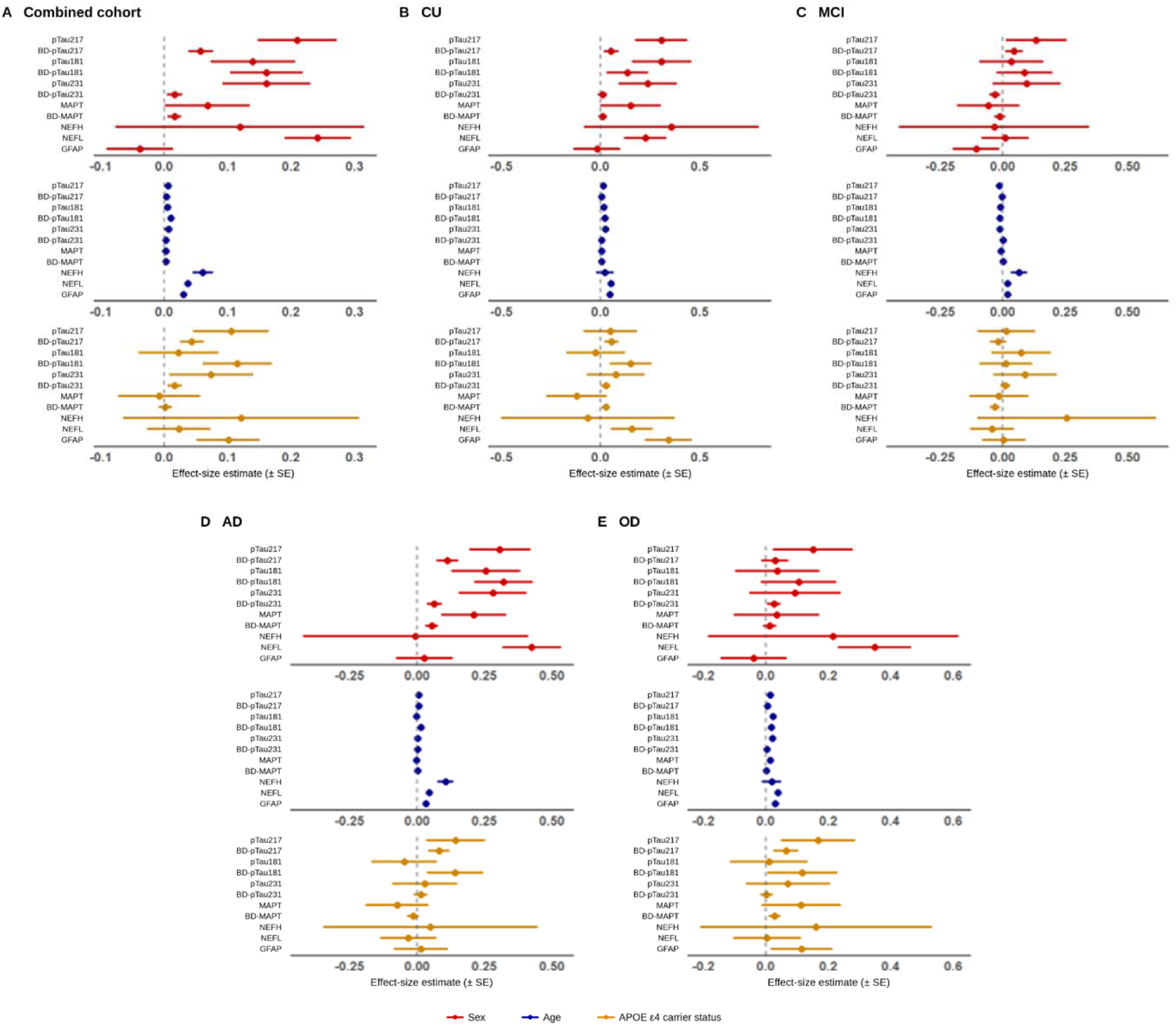
Associations of demographic variables with selected Alzheimer disease-related biomarkers. Effect estimates and standard errors are shown separately for sex, age, and APOE ε4 carrier status in a, the combined cohort; b, cognitively unimpaired participants; c, participants with mild cognitive impairment; d, participants with Alzheimer disease; and e, participants with other dementia. The other dementia group includes participants with mixed dementia and vascular dementia.

### Proteomic Profiles by Diagnostic Group

Figure 3 summarizes the direction, magnitude, and diagnostic-group overlap of biomarkers that differed significantly from CU after false discovery rate correction. Supplementary Figure 2 presents the corresponding volcano plots for MCI, AD, RD-M, and RD-V relative to CU, together with UpSet plots showing the overlap of upregulated and downregulated biomarkers across comparisons. AD had the largest number of biomarkers unique to a single diagnostic-group comparison, including 11 upregulated and 10 downregulated biomarkers. MME and UCHL1 were downregulated in AD, RD-M, and RD-V, but not MCI. MME showed the largest effect in each dementia-versus-CU comparison, making it the most consistent diagnostic-group signal in the panel. Several biomarkers overlapped across groups. IL5 was upregulated in MCI, AD, and RD-V; YWHAZ was upregulated in MCI, RD-V, and RD-M; and ENO2, PRDX6, and RUVBL2 were downregulated in both AD and RD-V.

**Figure 3.**
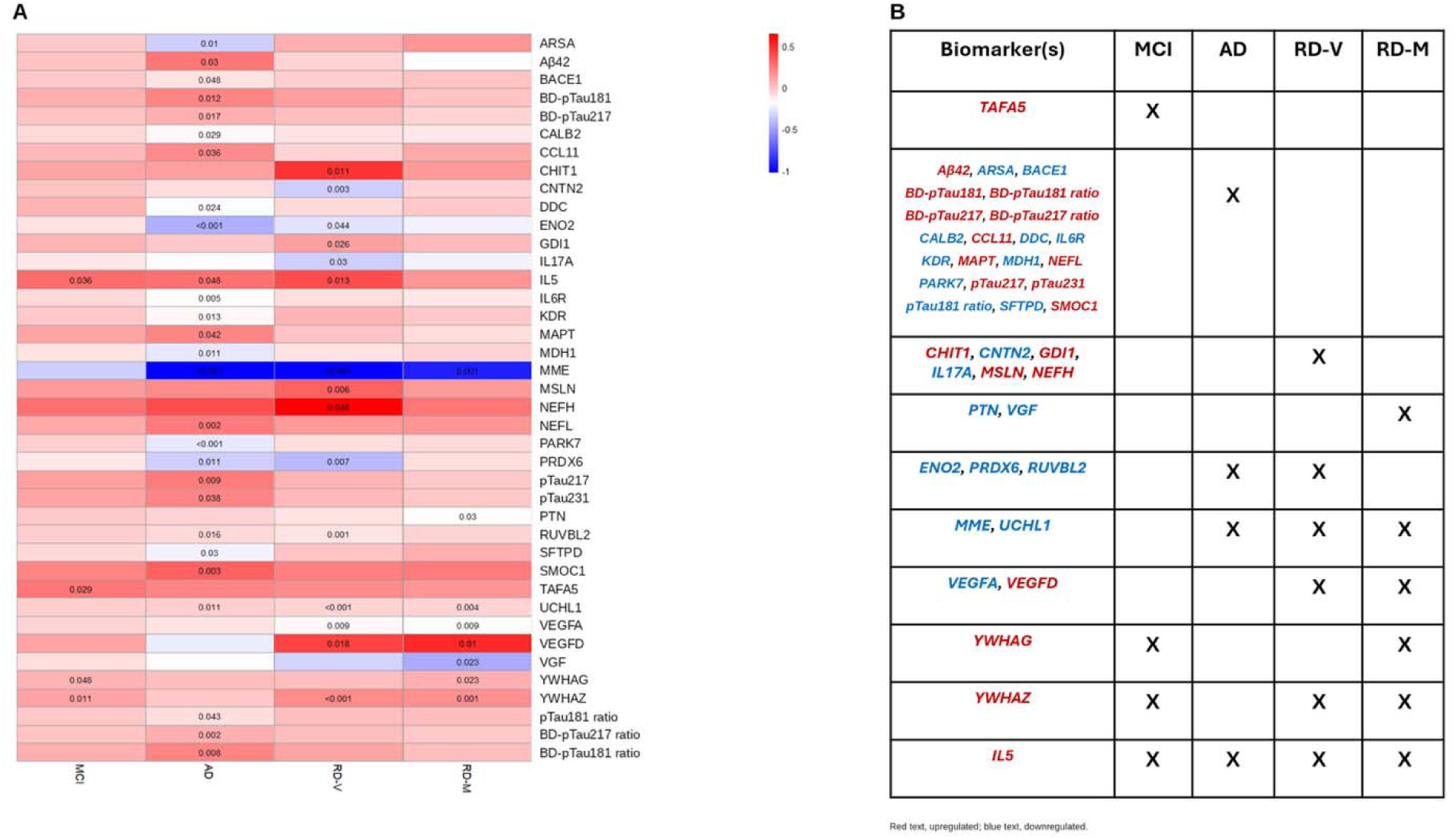
Plasma biomarkers differing from cognitively unimpaired participants by diagnostic group. a, Heatmap showing the direction and magnitude of the rank-regression coefficients for MCI, AD, RD-V, and RD-M relative to CU. Red indicates higher and blue indicates lower biomarker expression relative to CU. Values shown within cells indicate adjusted P values for comparisons meeting the false discovery rate-adjusted P < .05 threshold. b, Matrix identifying the diagnostic-group comparisons in which each biomarker remained significant after false discovery rate correction.

### Random Forest Classification

Using the 27 biomarkers that differed significantly between AD and CU, the random forest importance analysis identified SMOC1, NEFL, MME, BD-pTau217 ratio, and MDH1 as the five highest-ranked predictors (Figure 4b and Supplementary Figure 3). For CU-versus-AD classification, the highest mean cross-validated AUC was obtained using 10 biomarkers together with age, sex, and APOE ε4 carrier status (AUC = 0.768, SE = 0.0336). The five-biomarker minimum best model included SMOC1, NEFL, MME, BD-pTau217 ratio, and MDH1 and achieved an AUC of 0.738 (SE = 0.027; Figure 4a). By comparison, BD-pTau217 alone yielded an AUC of 0.612, indicating limited discrimination between the clinically adjudicated CU and AD groups.

**Figure 4.**
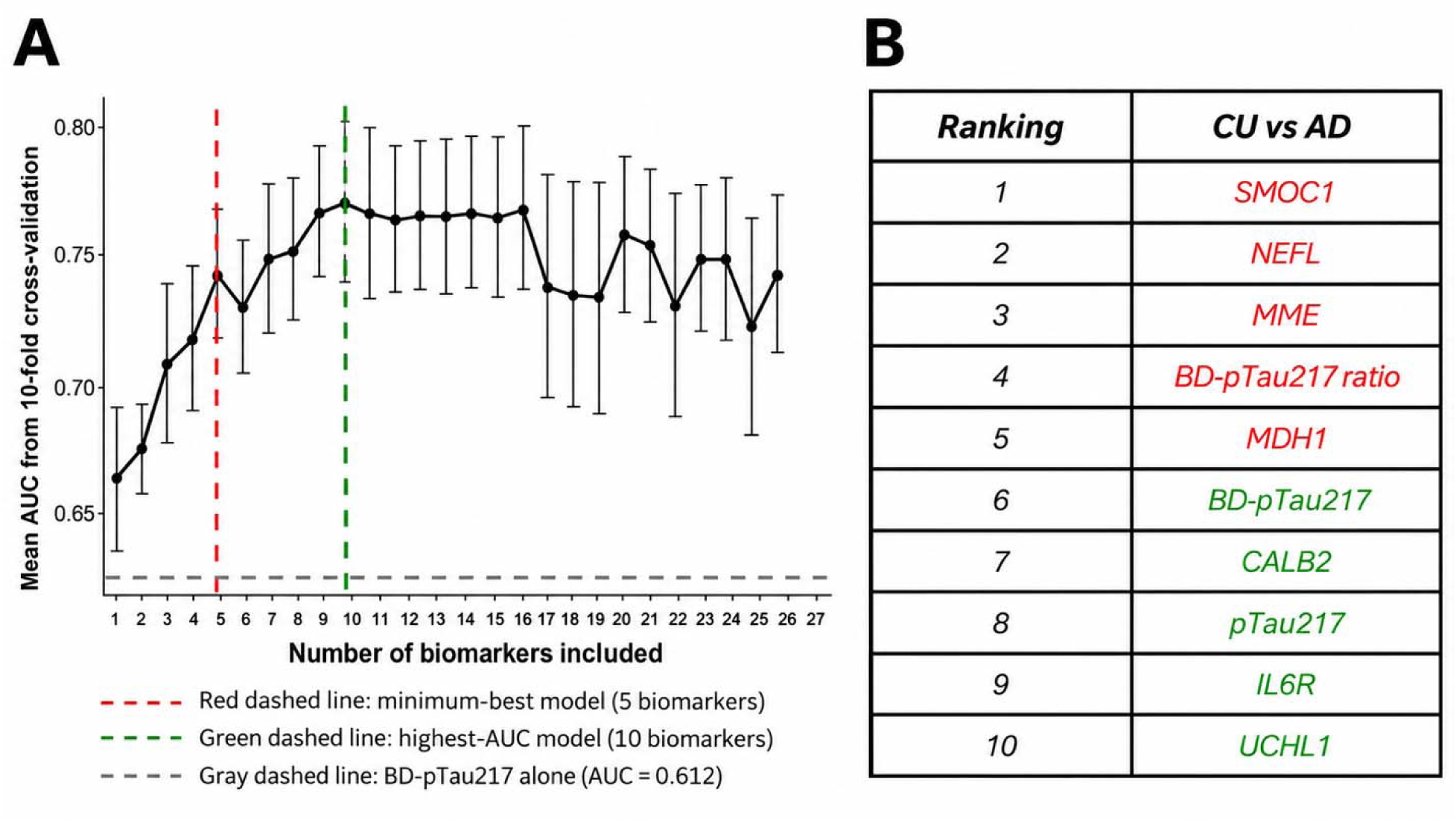
Random forest classification of cognitively unimpaired participants and participants with Alzheimer disease. a, Mean 10-fold cross-validated area under the receiver operating characteristic curve according to the number of biomarkers included in the model together with age, sex, and APOE ε4 carrier status. The red dashed line identifies the five-biomarker minimum best model, the green dashed line identifies the 10-biomarker model with the highest mean AUC, and the gray horizontal line indicates the AUC obtained using BD-pTau217 alone. b, The 10 highest-ranked biomarkers based on permutation importance from the initial random forest model. AUC, area under the receiver operating characteristic curve; CU, cognitively unimpaired; AD, Alzheimer disease. Error bars indicate standard errors across the 10 cross-validation folds. Red text identifies biomarkers in the five-biomarker minimum-best model, and green text identifies the additional biomarkers included in the highest-AUC 10-biomarker model.

The 27 candidate biomarkers were selected and initially ranked using the full dataset before cross-validation rather than separately within each training fold. The cross-validated AUC estimates should therefore be considered exploratory because this approach may have produced optimistic estimates of classification performance.

## DISCUSSION

To our knowledge, this is the first large NULISA-based proteomic study of cognitive aging and dementia in Central Africa. A recent Nigerian study used Simoa and NULISA to examine blood biomarkers in an indigenous African cohort,^18^ and our findings extend this work to older adults in Kinshasa. The results highlight the importance of demographic adjustment and population-specific validation when interpreting plasma proteomic profiles in this setting.

Sex was the strongest source of variation in protein levels, followed by age. Demographic and health factors, including age, sex, renal function, body mass index, and vascular comorbidity, can influence blood biomarker concentrations.^14,15^ In this Congolese cohort, the effects associated with sex were broader and larger than those associated with age, a pattern that should be replicated before population-specific reference ranges are established. The Nigerian VALIANT study also reported sex-related biomarker differences.^18^ APOE ε4 carrier status had a narrower association in the present study: after false discovery rate correction, only the pTau217 ratio, pTau231 ratio, BD-pTau217 ratio, and CRP differed by carrier status. These findings support demographic adjustment and locally validated reference values rather than direct application of thresholds developed in other populations.

AD showed the largest set of diagnosis-associated proteins, including markers related to amyloid and tau biology, neuroaxonal injury, and inflammation. GFAP was not significantly associated with AD in this cohort, in contrast to the Nigerian study,^18^ whereas CCL11, IL5, and IL6R differed in the AD comparison. Because infectious, inflammatory, nutritional, and cardiometabolic conditions were not modeled directly, these findings should not be attributed to any specific exposure. They instead identify pathways and covariates that should be examined in future longitudinal studies.

MME was downregulated in AD, mixed dementia, and vascular dementia and showed the largest effect in each dementia-versus-CU comparison, but it was not significant in MCI. MME encodes neprilysin, an enzyme involved in amyloid-β degradation,^22^ which makes this signal biologically relevant. However, plasma MME abundance does not establish neprilysin activity in the brain, and the cross-sectional association cannot determine directionality or causation. The finding is therefore best viewed as hypothesis-generating and requires replication with longitudinal data and measures that more directly assess central nervous system biology.

BD-pTau217 alone showed limited discrimination between clinically adjudicated CU and AD in this cohort, with an AUC of 0.612. Prior studies have generally evaluated BD-pTau217 against amyloid PET or other biological reference standards,^10,11^ whereas the present outcome was a clinical diagnosis based on cognitive, functional, and neurological findings. These results therefore do not establish that BD-pTau217 lacks value in Congolese adults. Rather, they show that performance against a clinically defined dementia diagnosis may differ from performance against biomarker-confirmed AD pathology. Studies incorporating amyloid PET, cerebrospinal fluid biomarkers, or other biological reference standards are needed to evaluate BD-pTau217 in this population.

Multianalyte models performed better than BD-pTau217 alone for classifying CU and AD, consistent with work suggesting that broader panels may capture disease heterogeneity. These estimates nevertheless require cautious interpretation. Candidate biomarkers were selected and initially ranked using the full dataset before cross-validation rather than separately within each training fold, which may have produced optimistic AUC estimates. External validation and fully nested feature selection are therefore needed before these models are considered for clinical use.

Strengths of this study include the large, well-characterized Central African cohort, multidisciplinary clinical adjudication, and use of a high-sensitivity proteomic platform. Limitations include the cross-sectional design, multiple-testing burden, exclusion of the RD-P and RD-F groups because of their small sample sizes, inference of APOE ε4 carrier status through proteotyping rather than direct genotyping, and absence of amyloid PET or cerebrospinal fluid confirmation. Candidate biomarker selection and importance ranking were performed using the full dataset before cross-validation, which may have inflated classification performance. We also could not evaluate several factors that may affect circulating proteins in this setting, including chronic infections, inflammatory disorders, nutritional deficiencies, thyroid disease, hemolytic conditions, and vascular or metabolic comorbidity. The classification findings should therefore be considered exploratory.

Overall, sex and age accounted for substantial plasma protein variation, MME provided a consistent hypothesis-generating signal across the dementia groups, and BD-pTau217 alone showed limited discrimination between clinically adjudicated CU and AD. These findings support demographic adjustment, population-specific validation, and biological confirmation of blood-based biomarkers before clinical thresholds developed elsewhere are applied in Central African populations.

## Methods

### Participants

The EVCD-RDC is a prospective longitudinal cohort based in Kinshasa, DRC. Eligibility criteria included Congolese citizenship and residence in the DRC; age 65 years or older; a valid home address and telephone number; contact information for a caregiver or informant; willingness to participate in baseline and follow-up assessments; the ability to provide informed consent independently or, when applicable, through a legally authorized representative; fluency in Lingala or French; and no history of a neurodevelopmental, psychiatric, or neurological disorder affecting the central nervous system. Individuals with significant visual or hearing impairment were excluded.

### Ethics approval and consent

The study was approved by the Research Ethics Committee of the School of Public Health at the University of Kinshasa (initial approval no. ESP/CE/054c/2024; renewal no. ESP/CE/054D/2026) and the Emory University Institutional Review Board (IRB00113463; modification MOD006). All participants, or their legally authorized representatives when applicable, provided written informed consent before participation.

### Clinical and Cognitive Assessment

Participants completed measures from the Uniform Data Set version 3, screening instruments, the African Neuropsychology Battery Short Version, intelligence tests, functional measures, and other questionnaires. Measures were selected to support harmonization with international dementia cohorts. All instruments were culturally adapted, translated into French and Lingala, and piloted before use. The neurological examination assessed cranial nerves, gait and balance, coordination, reflexes, muscle strength and tone, and sensation. Clinical findings were used to identify neurological conditions that could contribute to cognitive impairment and to inform differential diagnosis.

### Clinical Adjudication

A multidisciplinary team comprising a neuropsychologist (JI), neurologists (EE and PJ), a psychiatrist (GG), and an internist (TT) classified participants as CU, MCI, suspected AD, or another dementia subtype. Adjudication proceeded in two stages: determination of cognitive status and assignment of the most likely etiology based on the cognitive profile, functional status, medical history, neurological findings, and course of symptoms. Participants with no impaired cognitive domains and intact functioning, including an Identification and Intervention for Dementia in Elderly Africans (IDEA) cognitive screen score of 30 or higher, were classified as CU. Impairment in one cognitive domain with preserved daily functioning was classified as MCI. Impairment in at least two domains, defined as performance more than 1.5 standard deviations below the reference mean, together with functional decline and a screening score below 30, was classified as dementia. A progressive, memory-predominant course supported suspected AD, whereas a stepwise or fluctuating course, prominent medical contributors, or marked personality change supported classification as another dementia subtype.

Demographically adjusted predicted scores were generated with linear regression models for the screening, cognitive, intelligence, and functional measures. Age, years of education, and sex were entered as predictors of cognitive test scores, and the magnitude and statistical significance of their associations were examined within the Congolese clinical context. Cognitive impairment was quantified using z scores derived from CU reference data from the prior study. For each participant, the z score was calculated by subtracting the CU reference mean from the participant’s predicted score and dividing the difference by the CU reference standard deviation.

### Biospecimen Collection

Trained laboratory technicians collected biospecimens at the Centre Médical de Kinshasa according to international collection standards. Blood was collected in one 10-mL tube without anticoagulant for serum, one 10-mL EDTA tube for plasma, and two 4-mL EDTA tubes for DNA. Samples collected without anticoagulant remained at room temperature for at least 30 minutes before processing. Blood was centrifuged at 4,000 rpm for 10 minutes, and serum and plasma were transferred into sterile 0.5-mL aliquots. Plasma was stored initially at −20 °C and subsequently at −80 °C. The broader protocol also collected PAXgene RNA, urine, saliva, and fecal specimens.

### APOE ε4 Proteotyping

APOE ε4 carrier status was inferred through APOE proteotyping using APOE4 normalized protein quantification (NPQ) values generated by the NULISAseq CNS Disease Panel 120. Participants with APOE4 NPQ values below 5.78 were classified as noncarriers, whereas those with values of 5.78 or higher were classified as carriers. Heterozygous and homozygous APOE ε4 carrier status was not differentiated because the proteotyping approach was less accurate for distinguishing these groups.^23^

### NULISA Biomarker Measurement

Plasma proteins were measured with the NULISAseq CNS Disease Panel 120 (Alamar Biosciences, Inc.) on an ARGO HT system according to published protocols. Thawed plasma was centrifuged at 10,000 × g for 10 minutes to remove particulates and then incubated with capture and detection antibodies linked to DNA barcodes. The assay targeted 131 protein biomarkers across biological processes implicated in neurodegenerative disease and included mCherry as an internal standard. After immunocomplex purification, T4 DNA ligase and a specific ligator sequence joined the capture and detection barcodes to form reporter DNA, which was quantified by next-generation sequencing. Sequencing counts were normalized to the mCherry internal standard, harmonized with an interplate control, and log -transformed to generate NULISA Protein Quantification values. Samples were analyzed across 10 plates. The vendor-provided sample control was assayed in triplicate on each plate, and an in-house pooled plasma quality-control sample was assayed once per plate. Median intraplate and interplate coefficients of variation for the sample controls were 6.3% and 5.8%, respectively. The median interpolated interplate coefficient of variation for the three in-house quality-control samples was approximately 11%.

### Statistical Analysis

Analyses were conducted in R version 4.3.3 with RStudio (R Core Team, 2024). Packages included reshape2, ggplot2, dplyr, ranger, randomForest, caret, Rfit, UpSetR, and pROC. Only participants with corresponding biomarker data were included, and no biomarker measurements were missing because of analytical issues. One age value had been recorded incorrectly as the participant’s birth year and was corrected by recalculating age from the recorded birth year. There were no missing values for sex or diagnostic group. Participants classified as Parkinson disease-related dementia (RD-P) or frontotemporal dementia (RD-F) were excluded because each group included fewer than 10 participants.

### Rank-Based Regression

For each biomarker, rank-based regression estimated the associations of diagnostic group, age, sex, and APOE ε4 carrier status with protein expression. The model was specified as follows:

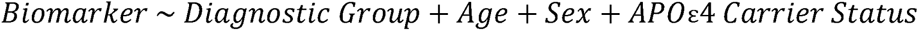

Regression coefficients served as effect estimates. P values were adjusted for multiple comparisons with the false discovery rate, and an adjusted P value below .05 was considered significant. CU, female sex, and APOE ε4 noncarriership were the reference categories for diagnostic group, sex, and APOE ε4 status, respectively. Age was modeled as a continuous variable.

### Random Forest Classification

Random forest models were used to identify biomarker combinations for classifying CU versus AD. The analysis proceeded in three steps. First, a classification random forest included the 27 biomarkers that differed significantly between AD and CU after adjustment for age, sex, and APOE ε4 carrier status. These biomarkers were selected using the full dataset. This initial model did not include demographic covariates. Biomarkers were ranked using permutation-based mean decrease in accuracy, also using the full dataset. The model used 1,000 trees, mtry = floor(√[number of variables − 1]), and permutation importance. Second, 27 classification random forest models were fitted using 10-fold cross-validation stratified by outcome group, with a random seed of 8888. For each k from 1 through 27, the model included the first k biomarkers in the importance ranking together with age, sex, and APOE ε4 carrier status. Mean area under the receiver operating characteristic curve (AUC) and its standard error were calculated across folds. Each model used 1,000 trees, mtry = floor(√k), and permutation importance. Biomarker selection and initial importance ranking were not repeated separately within each training fold. Third, the best model was defined as the model with the highest mean AUC. The minimum best model was defined as the model with the fewest biomarkers whose mean AUC fell within one standard error of the best model.

## Data Availability

All data produced in the present study are available upon reasonable request to the authors

## Data availability

De-identified data supporting the findings of this study are available from the corresponding author upon reasonable request, subject to applicable ethics approvals and data-use agreements.

## Code availability

The analysis code used in this study is available from the corresponding author upon reasonable request.

## Acknowledgments

This work was supported by an Alzheimer’s Association Research Grant (23AARG-1027059), the Emory Goizueta Alzheimer’s Disease Research Center, the National Institute on Aging of the National Institutes of Health (P30AG066511 and AG066511-02S1), and the National Center for Advancing Translational Sciences of the National Institutes of Health (UL1TR002378). The funders had no role in the study design, data collection, data analysis, interpretation of the findings, or preparation of the manuscript. The content is solely the responsibility of the authors and does not necessarily represent the official views of the National Institutes of Health. The authors thank the study participants and their families, the community partners who supported recruitment, and the clinical and research staff involved in the EVCD-RDC study and biological sample collection.

## Author contributions

J.I. conceived and supervised the study, obtained funding, oversaw cohort development and data collection, contributed to clinical adjudication, and revised the manuscript. S.S.P. contributed to the interpretation of the findings and drafting and revision of the manuscript. T.K. and T.B. contributed to the statistical methodology, formal analysis, interpretation of the findings, and revision of the manuscript. X.Z. contributed to statistical methodology and formal analysis. C.O., G.G., E.E., J.M.P., T.T., F.M.S., J.P.T., S.M., and L.M. contributed to participant recruitment, clinical assessment, data collection, and/or clinical adjudication. A.A. and A.G. contributed to manuscript revision. All authors reviewed and approved the final manuscript.

## Figure Legends

Supplementary Figure 1. Associations of age and APOE ε4 carrier status with biomarker expression. a, Volcano plot showing regression coefficients and P values for associations between age and biomarker expression. b, Age distributions for the three biomarkers with the largest positive associations (NEFH, NEFL, and CHI3L1) and the three biomarkers with the largest negative associations (MME, FCN2, and BD-pTau231). c, Volcano plot showing regression coefficients and P values for associations between APOE ε4 carrier status and biomarker expression. d, Distributions of pTau217 ratio, pTau231 ratio, BD-pTau217 ratio, and CRP by APOE ε4 carrier status. The dashed lines indicate the false discovery rate-adjusted P < .05 threshold.

Supplementary Figure 2. Biomarker expression by diagnostic group relative to cognitively unimpaired participants. Volcano plots compare a, mild cognitive impairment; b, Alzheimer disease; c, mixed dementia; and d, vascular dementia with the cognitively unimpaired group. Models were adjusted for age, sex, and APOE ε4 carrier status. Dashed lines indicate the false discovery rate-adjusted P < .05 threshold. Labeled biomarkers met the adjusted significance criterion. UpSet plots show biomarkers that were significantly e, upregulated and f, downregulated across diagnostic-group comparisons after false discovery rate correction.

Supplementary Figure 3. Biomarker selection and importance-ranking workflow. Twenty-seven biomarkers that differed significantly between participants with Alzheimer disease and cognitively unimpaired participants after adjustment for age, sex, and APOE ε4 carrier status were entered into a random forest classification model. Biomarkers were ranked using permutation-based mean decrease in accuracy. Candidate biomarker selection and initial importance ranking were performed using the full dataset before cross-validation.

